# Barriers and Facilitators to Participation in Physical Activity Programmes for Socially Isolated Older Adults: A Qualitative Systematic Review Protocol

**DOI:** 10.1101/2025.09.30.25337030

**Authors:** Ravi Shankar, Fiona Devi, Xu Qian

## Abstract

**Background:** Social isolation among older adults represents a significant public health challenge associated with reduced physical activity, functional decline, and increased mortality. Physical activity programmes offer potential interventions to address both isolation and physical deconditioning, yet participation rates remain suboptimal, particularly among socially isolated older adults who may face unique barriers to engagement.

**Objective:** To systematically identify, appraise, and synthesize qualitative evidence on barriers and facilitators to participation in physical activity programmes among socially isolated older adults, examining experiences, perceptions, and contextual factors that influence engagement across diverse settings and programme types.

**Methods:** This qualitative systematic review protocol follows the Enhancing Transparency in Reporting the Synthesis of Qualitative Research (ENTREQ) guidelines and PRISMA-P framework. Comprehensive searches will be conducted in MEDLINE, CINAHL, PsycINFO, Scopus, Web of Science, Embase, and AgeLine from inception to December 2025, without language restrictions. Grey literature will be searched through ProQuest Dissertations, OpenGrey, and relevant organizational websites. Two independent reviewers will screen studies using Covidence software, with eligibility criteria including qualitative or mixed-methods studies exploring participation experiences of socially isolated adults aged 60 and above in physical activity programmes. Quality assessment will employ the Critical Appraisal Skills Programme (CASP) Qualitative Checklist.

**Data Synthesis:** Thematic synthesis will be conducted following Thomas and Harden’s approach, involving line-by-line coding, development of descriptive themes, and generation of analytical themes. The Theoretical Domains Framework (TDF) will guide organization of barriers and facilitators across fourteen domains of behavior change. Confidence in review findings will be assessed using GRADE-CERQual.

**Expected Outcomes:** Comprehensive taxonomy of multi-level barriers and facilitators, development of a conceptual model linking social isolation to physical activity participation, identification of intervention components addressing specific barriers, and evidence-based recommendations for programme design and implementation strategies tailored to socially isolated older adults.

## Introduction

Social isolation among older adults has emerged as a critical public health concern, with prevalence estimates suggesting that between 20% to 34% of community-dwelling older adults experience significant social isolation, a figure that has been substantially exacerbated by the COVID-19 pandemic and associated public health restrictions [1]. The intersection of social isolation and physical inactivity creates a particularly concerning cycle, as socially isolated older adults are significantly less likely to engage in regular physical activity, while physical inactivity further limits opportunities for social connection and community engagement [2]. This bidirectional relationship underscores the potential value of physical activity programmes that explicitly address both physical and social dimensions of health, yet evidence suggests that socially isolated older adults face unique challenges in accessing and maintaining participation in such programmes [3-5].

The conceptualization of social isolation encompasses both objective and subjective dimensions, with objective isolation referring to quantifiable aspects such as living alone, limited social network size, and infrequent social contact, while subjective isolation relates to perceived loneliness and feelings of disconnection from others [6]. This multidimensional nature of social isolation has important implications for understanding participation in physical activity programmes, as individuals may experience different types and degrees of isolation that influence their capacity and motivation to engage in group-based or community activities [7]. Furthermore, the pathway from social isolation to physical activity participation is mediated by complex psychological, social, and environmental factors that require systematic examination to inform effective intervention strategies [8].

Physical activity programmes for older adults have evolved considerably over recent decades, encompassing diverse formats ranging from structured exercise classes and fitness center-based programmes to community walking groups, tai chi sessions, and technology-supported home-based interventions [9-11]. While substantial evidence demonstrates the physical and psychosocial benefits of regular physical activity for older adults, including improved functional capacity, reduced fall risk, enhanced mood, and increased social interaction, participation rates remain disappointingly low, particularly among vulnerable populations such as socially isolated individuals [12]. Understanding the specific barriers and facilitators that influence participation among socially isolated older adults is essential for developing targeted strategies to improve programme reach and effectiveness.

The theoretical understanding of physical activity behavior in older adults has been informed by multiple behavioral theories, including Social Cognitive Theory, the Theory of Planned Behavior, and the Social Ecological Model, each offering valuable perspectives on the multi-level factors influencing participation [13]. However, the unique circumstances of socially isolated older adults suggest that traditional theoretical frameworks may require adaptation or integration to fully capture the complex interplay between social isolation, physical capacity, psychological factors, and environmental constraints that shape participation decisions [14]. The development of more nuanced theoretical models specific to this population requires systematic synthesis of qualitative evidence that captures the lived experiences and perspectives of socially isolated older adults themselves.

Recent systematic reviews have examined various aspects of physical activity participation among older adults, including barriers and facilitators in general older adult populations, the effectiveness of different intervention approaches, and the role of social support in promoting physical activity [15, 16]. However, no systematic review has specifically focused on the unique experiences and perspectives of socially isolated older adults regarding participation in physical activity programmes. This gap is particularly significant given growing recognition that socially isolated individuals may face distinct challenges that are not adequately addressed by generic physical activity promotion strategies designed for the broader older adult population [17].

### Background and Rationale

The demographic transition toward population aging represents one of the most significant global challenges of the 21st century, with the number of adults aged 65 and older expected to double from 727 million in 2020 to 1.5 billion by 2050 [18]. This demographic shift is accompanied by increasing prevalence of social isolation, driven by factors including changing family structures, urbanization, increased mobility, and the loss of social roles and relationships that often accompanies aging [19]. The health consequences of social isolation are profound and well-documented, with meta-analytic evidence demonstrating that social isolation increases mortality risk by 29%, comparable to well-established risk factors such as obesity and physical inactivity [20].

The relationship between social isolation and physical inactivity is particularly concerning from a public health perspective, as both conditions independently contribute to adverse health outcomes while potentially reinforcing each other through various biological, psychological, and social pathways [21]. Socially isolated older adults demonstrate lower levels of physical activity across multiple domains including leisure-time activity, transportation-related activity, and household activities, with longitudinal studies suggesting that social isolation predicts subsequent decline in physical activity levels independent of baseline health status and demographic factors [22]. The mechanisms underlying this relationship are multifaceted, including reduced motivation for self-care, limited social support for activity participation, decreased access to activity opportunities, and the absence of social modeling and reinforcement that facilitate behavior change and maintenance [23].

Physical activity programmes designed for older adults have proliferated in recent years, reflecting growing awareness of the importance of physical activity for healthy aging and the specific exercise needs and preferences of older populations [24]. These programmes vary widely in format, intensity, setting, and delivery mode, ranging from high-intensity functional training in fitness facilities to gentle chair-based exercises in residential care settings, and from instructor-led group classes to self-directed home programmes supported by digital technologies [25]. While this diversity potentially allows for tailoring to individual preferences and capabilities, it also creates challenges in identifying which programme characteristics are most suitable for engaging socially isolated older adults who may have limited experience with formal exercise programmes and face multiple barriers to participation [26].

The evidence base for physical activity interventions specifically targeting socially isolated older adults remains limited and fragmented, with most studies focusing on general older adult populations or specific clinical groups without explicit consideration of social isolation status [27]. Existing interventions that have included socially isolated participants suggest that standard approaches may be insufficient, with higher dropout rates, lower adherence, and reduced effectiveness observed among socially isolated individuals compared to their more socially connected counterparts [17]. These findings highlight the need for targeted strategies that address the unique needs and circumstances of socially isolated older adults, yet the development of such strategies is hampered by limited understanding of the specific factors that influence participation decisions in this population.

Qualitative research offers particular value in understanding the complex, contextual, and subjective factors that influence physical activity participation among socially isolated older adults, providing insights into lived experiences, meaning-making processes, and the nuanced interplay between individual, social, and environmental factors [28]. Individual qualitative studies have identified various themes related to participation barriers and facilitators, including the role of past experiences with physical activity, the importance of programme atmosphere and social dynamics, practical considerations such as transportation and scheduling, and the influence of health professionals and family members on participation decisions [16]. However, these individual studies are limited in scope and generalizability, and no systematic synthesis has been conducted to integrate findings across studies and contexts.

The theoretical landscape for understanding health behavior in older adults has evolved to recognize the importance of social and environmental factors alongside individual-level determinants, with frameworks such as the Social Ecological Model emphasizing the multi-level influences on behavior [29]. The Theoretical Domains Framework (TDF), which integrates constructs from multiple behavior change theories into fourteen domains, provides a comprehensive structure for organizing and understanding the diverse factors that influence behavior change, including knowledge, skills, social influences, environmental context, and behavioral regulation [30]. Application of the TDF to physical activity participation among socially isolated older adults can facilitate systematic identification of intervention targets and the development of theory-informed strategies to address specific barriers and leverage facilitators.

The current policy context increasingly recognizes the importance of addressing social isolation and promoting physical activity among older adults, with initiatives such as the WHO’s Global Action Plan on Physical Activity 2018-2030 and various national strategies targeting these interconnected challenges [31]. However, translation of policy objectives into effective programmes requires detailed understanding of implementation challenges and opportunities at the community level, particularly for reaching and engaging marginalized populations such as socially isolated older adults [32]. Qualitative evidence synthesis can inform policy and practice by identifying common implementation challenges across contexts, successful strategies for overcoming barriers, and key programme features that facilitate engagement of socially isolated participants.

### Theoretical Framework

This systematic review will employ the Theoretical Domains Framework (TDF) as the primary organizing structure for understanding barriers and facilitators to physical activity participation among socially isolated older adults [33]. The TDF provides a comprehensive, theory-informed approach to identifying factors influencing behavior change across fourteen domains: knowledge, skills, social/professional role and identity, beliefs about capabilities, optimism, beliefs about consequences, reinforcement, intentions, goals, memory/attention/decision processes, environmental context and resources, social influences, emotion, and behavioral regulation [34]. This framework has been successfully applied to understanding physical activity behavior in various populations and offers particular value for systematically organizing the diverse factors that may influence participation among socially isolated older adults.

The knowledge domain encompasses awareness and understanding of physical activity benefits, programme availability, and participation requirements, which may be particularly limited among socially isolated older adults who have reduced access to health information through social networks [35]. The skills domain includes both physical capabilities required for programme participation and social skills necessary for group interaction, with social isolation potentially affecting both dimensions through physical deconditioning and reduced opportunities for social skill maintenance [33]. The social/professional role and identity domain relates to how individuals perceive themselves in relation to physical activity and social participation, with social isolation potentially disrupting identity formation around active aging and community engagement [28].

Beliefs about capabilities, closely aligned with self-efficacy concepts from Social Cognitive Theory, may be particularly compromised among socially isolated older adults who lack social support and vicarious learning opportunities that build confidence in physical activity participation [13]. The optimism domain captures general positive expectations about outcomes, while beliefs about consequences encompasses specific anticipated outcomes of participation, both of which may be influenced by the negative cognitive patterns often associated with social isolation [7]. The reinforcement domain addresses rewards and incentives for participation, which may be limited for socially isolated individuals who lack social recognition and support for their efforts.

The intentions and goals domains capture motivational aspects of behavior, with social isolation potentially affecting both the formation of participation intentions and the establishment of meaningful physical activity goals [36]. Memory, attention, and decision processes become increasingly important considerations for older adults, with social isolation potentially exacerbating cognitive challenges through reduced cognitive stimulation and increased stress [37]. Environmental context and resources encompasses the physical, social, and organizational factors that enable or constrain participation, with socially isolated individuals potentially facing multiple environmental barriers including transportation challenges, financial constraints, and limited awareness of available programmes.

Social influences represent a particularly critical domain for socially isolated older adults, as the absence of social support, role models, and social norms promoting physical activity may fundamentally undermine participation motivation and capability [8]. The emotion domain captures affective responses including fear, anxiety, and depression that may both result from and contribute to social isolation while creating additional barriers to programme participation [17]. Finally, behavioral regulation involves self-monitoring, planning, and other self-regulatory strategies that may be compromised in socially isolated individuals who lack external structure and support for behavior change.

Integration of the TDF with social isolation-specific considerations suggests several unique pathways through which isolation may influence physical activity participation. Social isolation may create cascading effects across multiple TDF domains, with reduced social influences leading to diminished beliefs about capabilities, limited reinforcement opportunities, and compromised behavioral regulation strategies [14]. Additionally, the bidirectional relationship between social isolation and physical inactivity suggests that interventions must address both social and physical activity goals simultaneously, requiring careful consideration of how programme design features can facilitate social connection while promoting physical activity engagement.

### Objectives

This systematic review aims to comprehensively identify, critically appraise, and synthesize qualitative evidence on the barriers and facilitators influencing participation in physical activity programmes among socially isolated older adults, with the overarching goal of informing the development of targeted interventions and implementation strategies that effectively engage this vulnerable population. The primary objective encompasses exploring the lived experiences, perceptions, and decision-making processes of socially isolated older adults regarding physical activity programme participation, examining how social isolation intersects with other individual, social, and environmental factors to shape engagement patterns, and understanding how different programme characteristics and delivery approaches influence accessibility and acceptability for socially isolated individuals. This comprehensive examination will utilize the Theoretical Domains Framework to systematically organize findings across multiple behavioral domains, enabling identification of key intervention targets and mechanisms of action. The review will specifically investigate how social isolation influences each TDF domain, examining unique patterns and pathways that distinguish socially isolated older adults from the general older adult population, while also exploring how programme features can address isolation-specific barriers and leverage facilitators to promote engagement. Through synthesis of diverse qualitative evidence from multiple contexts and programme types, the review aims to develop a nuanced understanding of the complex factors influencing participation decisions, identify successful strategies for overcoming barriers, and generate practical recommendations for programme design, implementation, and evaluation that are grounded in the perspectives and experiences of socially isolated older adults themselves.

## Methods

### Study Design

This qualitative systematic review will follow the Enhancing Transparency in Reporting the Synthesis of Qualitative Research (ENTREQ) guidelines to ensure comprehensive and transparent reporting of the review process and findings [38]. The protocol has been developed in accordance with the Preferred Reporting Items for Systematic Review and Meta-Analysis Protocols (PRISMA-P) statement, adapted for qualitative synthesis [39]. The review will employ a thematic synthesis approach as described by Thomas and Harden (2008) [40], which provides a structured method for integrating findings from multiple qualitative studies while preserving the richness and complexity of the original data. This approach is particularly suited to addressing questions about barriers and facilitators, as it enables both aggregation of findings across studies and generation of new interpretive insights that extend beyond the primary research.

The epistemological stance underpinning this review aligns with critical realism, acknowledging that while there are real barriers and facilitators to physical activity participation, our understanding of these factors is mediated through the interpretive lenses of research participants, primary researchers, and review authors [41]. This philosophical position supports the synthesis of findings from studies employing diverse qualitative methodologies while recognizing that different methodological approaches may reveal different aspects of the phenomenon under investigation. The review will maintain reflexivity throughout the synthesis process, with team members documenting their assumptions, decisions, and interpretations to ensure transparency and enable critical evaluation of the review findings.

### Search Strategy

A comprehensive search strategy has been developed in consultation with an information specialist experienced in systematic review methodology, utilizing both controlled vocabulary and free-text terms to maximize sensitivity while maintaining feasibility. The strategy combines four concept blocks: (1) older adults and related terms, (2) social isolation and loneliness concepts, (3) physical activity and exercise programmes, and (4) qualitative research designs and methods. These blocks will be combined using Boolean operators, with the search strategy adapted for each database according to specific indexing terms and syntax requirements.

The following electronic databases will be searched from inception to December 2025: MEDLINE (via PubMed), CINAHL (Cumulative Index to Nursing and Allied Health Literature), PsycINFO, Scopus, Web of Science Core Collection, Embase, and AgeLine. The selection of databases ensures comprehensive coverage of health sciences, psychology, social sciences, and gerontology literature. No language restrictions will be applied initially, with translation services utilized for potentially relevant non-English publications. Grey literature will be searched through ProQuest Dissertations and Theses Global, OpenGrey, and websites of relevant organizations including WHO, Age UK, National Institute on Aging, and International Association of Gerontology and Geriatrics.

The search terms for the older adults concept include: “older adult*,” “older people,” “elderly,” “senior*,” “aging,” “ageing,” “geriatric*,” and related MeSH/thesaurus terms. Social isolation terms encompass: “social isolation,” “socially isolated,” “loneliness,” “lonely,” “social disconnection,” “social exclusion,” “living alone,” “homebound,” “shut-in,” and associated controlled vocabulary. Physical activity programme terms include: “physical activity,” “exercise,” “physical fitness,” “exercise program*,” “fitness class*,” “group exercise,” “exercise intervention,” “movement program*,” “activity program*,” and related indexed terms. Qualitative research terms comprise: “qualitative,” “interview*,” “focus group*,” “ethnograph*,” “phenomenolog*,” “grounded theory,” “thematic analysis,” “content analysis,” “narrative,” “experience*,” “perception*,” “barrier*,” “facilitator*,” “enabler*,” and corresponding methodology filters.

Reference lists of included studies and relevant systematic reviews will be hand-searched to identify additional eligible studies. Forward citation searching will be conducted using Google Scholar and Web of Science for all included studies. Content experts in gerontology, physical activity, and social isolation will be contacted to identify unpublished or in-progress studies. Conference proceedings from the past five years of major gerontology and physical activity conferences will be searched, including the Gerontological Society of America, International Association of Gerontology and Geriatrics, and International Society of Physical Activity and Health.

### Eligibility Criteria

Studies will be included if they meet all of the following criteria: (1) include participants aged 60 years or older who are identified as socially isolated or lonely through either objective measures (living alone, limited social contact, small social network) or subjective assessment (self-reported loneliness, perceived isolation); (2) explore experiences, perceptions, barriers, or facilitators related to participation in structured physical activity or exercise programmes, including group classes, individual training, home-based programmes, or community-based activities; (3) employ qualitative data collection methods including interviews, focus groups, participant observation, or other recognized qualitative approaches; (4) present qualitative findings related to participation factors, even if embedded within mixed-methods studies; and (5) are published as peer-reviewed articles, dissertations, or research reports with sufficient methodological detail for quality assessment.

Studies will be excluded if they: (1) focus exclusively on physical activity behavior in general without specific attention to structured programmes or interventions; (2) examine only clinical populations with specific diseases unless social isolation is explicitly addressed as a key characteristic; (3) include only quantitative data without qualitative findings; (4) are intervention studies that report only outcome data without exploring participant experiences; (5) focus solely on care home or institutional settings without relevance to community-dwelling older adults; (6) are published only as conference abstracts, editorials, opinion pieces, or protocols without results; or (7) examine only professional or caregiver perspectives without including older adults’ own voices.

The definition of social isolation will be intentionally broad to capture diverse conceptualizations across studies, recognizing that isolation may be operationalized differently across cultural contexts and research traditions. Studies that focus on loneliness as the primary construct will be included given the conceptual overlap and frequent co-occurrence with social isolation. Similarly, physical activity programmes will be broadly defined to include various formats and delivery modes, from formal exercise classes to informal walking groups, provided they involve some degree of structure or organization beyond spontaneous individual activity.

### Study Selection

All identified citations will be imported into Covidence systematic review software for deduplication and screening management. The screening process will proceed in two stages, with two independent reviewers assessing studies at each stage. Prior to formal screening, a calibration exercise will be conducted using a random sample of 50 citations to ensure consistency in applying eligibility criteria, with discrepancies discussed and criteria clarified as needed to achieve acceptable inter-rater agreement (kappa ≥ 0.80).

Title and abstract screening will be conducted independently by two reviewers using standardized screening forms in Covidence. Studies will be retained for full-text review if they potentially meet inclusion criteria or if insufficient information is available to make a determination. Disagreements will be resolved through discussion, with a third reviewer consulted if consensus cannot be reached. Reasons for exclusion will be documented at the abstract screening stage to inform refinement of the search strategy if needed.

Full-text screening will follow the same independent dual-review process, with reviewers documenting specific reasons for exclusion using Covidence’s standardized categories aligned with the eligibility criteria. Where full texts are unavailable through institutional access or interlibrary loan, corresponding authors will be contacted with a request for manuscripts. For studies published in languages other than English, initial screening will be conducted using English abstracts where available, with full translation arranged for potentially eligible studies. A PRISMA flow diagram will be generated automatically through Covidence to document the study selection process.

### Data Extraction

A standardized data extraction form will be developed and piloted on three included studies to ensure comprehensive capture of relevant study characteristics and findings. The form will be refined based on pilot testing and team discussion before full implementation. Data extraction will be conducted independently by two reviewers, with discrepancies resolved through discussion and reference to original texts. The extraction form will be implemented in Covidence or a structured spreadsheet to facilitate data management and synthesis.

Study characteristics to be extracted include: author(s) and publication year; country and setting; study aims and research questions; theoretical or conceptual framework; methodology and epistemological approach; sampling strategy and sample size; participant characteristics including age, gender, living situation, and measures of social isolation; programme characteristics including type, format, duration, frequency, and delivery mode; data collection methods and procedures; data analysis approach; researcher reflexivity and positioning; and funding sources and potential conflicts of interest.

Findings related to barriers and facilitators will be extracted verbatim, including participant quotes, authors’ interpretations, and thematic categories presented in the primary studies. Each finding will be coded according to the relevant TDF domain(s) and classified as a barrier, facilitator, or both depending on context. Contextual information that may influence transferability of findings will be captured, including cultural factors, healthcare system characteristics, and programme-specific features that may moderate the influence of identified barriers and facilitators.

### Quality Assessment

Methodological quality will be assessed using the Critical Appraisal Skills Programme (CASP) Qualitative Checklist, a widely-used tool comprising ten questions addressing key aspects of qualitative research quality [42]. The CASP checklist examines: clarity of research aims; appropriateness of qualitative methodology; research design suitability; recruitment strategy; data collection methods; researcher-participant relationship; ethical considerations; rigor of data analysis; clarity of findings presentation; and research value. Each criterion will be assessed as “yes,” “no,” or “can’t tell,” with explanatory comments documenting the rationale for each assessment.

Two reviewers will independently assess the quality of each included study, with disagreements resolved through discussion and, if necessary, consultation with a third reviewer. Quality assessment will not be used to exclude studies but rather to provide context for interpretation of findings and assessment of confidence in review findings. A sensitivity analysis will explore whether excluding lower-quality studies substantially alters the review findings. Studies will be categorized as high, moderate, or low quality based on the number of criteria met, with particular weight given to clarity of findings, rigor of analysis, and consideration of researcher reflexivity.

The quality assessment will inform the GRADE-CERQual (Confidence in Evidence from Reviews of Qualitative research) assessment, which will be conducted for each review finding to indicate confidence in the synthesized evidence [43]. GRADE-CERQual assessment considers four components: methodological limitations of included studies; coherence of the review finding; adequacy of data contributing to the finding; and relevance of included studies to the review question. Each review finding will be assigned a confidence rating of high, moderate, low, or very low, with explanations provided for any downgrading decisions.

### Data Synthesis

Thematic synthesis will be conducted following the three-stage approach described by Thomas and Harden (2008): line-by-line coding of findings from primary studies, development of descriptive themes, and generation of analytical themes. This approach enables both aggregation of findings across studies and development of new interpretive insights that extend beyond the primary research. NVivo software will be used to manage the coding and synthesis process, facilitating systematic organization of data and maintenance of an audit trail documenting analytical decisions.

The first stage involves line-by-line coding of all text labeled as “results” or “findings” in included studies, including participant quotes and authors’ interpretations. Two reviewers will independently code a subset of studies to develop an initial coding framework, which will be refined through discussion and applied to remaining studies. Codes will be both inductive, emerging from the data, and deductive, informed by the TDF domains. New codes will be added iteratively as needed, with the coding framework regularly reviewed and refined to ensure consistency and comprehensiveness.

The second stage involves organizing codes into descriptive themes that remain close to the primary study findings. Descriptive themes will be developed through examining relationships between codes, identifying patterns across studies, and grouping related concepts. Each descriptive theme will be defined and illustrated with supporting data from multiple studies where available. The descriptive themes will be mapped to TDF domains while remaining open to themes that may not align with the framework, ensuring that important findings are not overlooked due to framework constraints.

The third stage involves developing analytical themes that go beyond the primary studies to generate new interpretive explanations and theoretical insights. This stage involves examining relationships between descriptive themes, identifying overarching patterns, and developing explanatory models that account for variation in findings across contexts. The analytical themes will address the review questions while generating new understandings of how social isolation influences physical activity participation through multiple pathways and mechanisms. A conceptual model will be developed to illustrate relationships between themes and their influence on participation outcomes.

## Discussion

This qualitative systematic review of barriers and facilitators to physical activity programme participation among socially isolated older adults addresses a critical gap in the evidence base for developing targeted interventions for this vulnerable population. The synthesis of qualitative evidence will provide nuanced understanding of the complex interplay between social isolation and physical activity participation, revealing pathways and mechanisms that are not captured through quantitative research alone. The application of the Theoretical Domains Framework ensures systematic examination of multiple behavioral influences while allowing for emergence of isolation-specific factors that may not be adequately captured by existing theoretical models. The review findings will have important implications for programme design, implementation strategies, and policy development aimed at promoting physical activity and reducing social isolation among older adults.

The theoretical contributions of this review extend beyond identifying specific barriers and facilitators to advancing conceptual understanding of how social isolation shapes health behavior in later life. By examining physical activity participation through the lens of social isolation, the review will illuminate how the absence or limitation of social connections creates cascading effects across multiple behavioral domains, from undermining self-efficacy and motivation to limiting access to resources and opportunities for engagement. The synthesis will contribute to theoretical development by identifying mechanisms through which social isolation influences behavior, patterns of interaction between different barriers and facilitators, and contextual factors that moderate these relationships. These theoretical insights will inform future research and intervention development by identifying key leverage points for behavior change and suggesting novel intervention strategies that address the unique needs of socially isolated older adults.

The practical implications for programme design and delivery are substantial, as the review will provide evidence-based guidance for developing physical activity programmes that are accessible, acceptable, and effective for socially isolated older adults. Understanding the specific barriers faced by this population will enable programme developers to proactively address challenges such as transportation difficulties, social anxiety, lack of confidence, and unfamiliarity with exercise environments. Similarly, identification of facilitators will highlight programme features and implementation strategies that successfully engage socially isolated participants, such as peer support mechanisms, graduated entry approaches, and explicit attention to social relationship building alongside physical activity goals. The review will also identify successful strategies for reaching and recruiting socially isolated older adults, who may be invisible to traditional recruitment approaches that rely on existing social networks and community connections.

The policy implications of this review relate to the broader public health agenda of promoting healthy aging and addressing social determinants of health. The findings will inform policy development by identifying system-level barriers that require policy intervention, such as transportation infrastructure, programme funding mechanisms, and professional training requirements. The review will also contribute to implementation science by identifying factors that influence successful translation of policy objectives into effective community-level programmes. By highlighting the intersection between social isolation and physical inactivity, the review supports integrated policy approaches that address multiple risk factors simultaneously rather than treating them as separate issues requiring distinct interventions.

The expected outcomes of this systematic review include development of a comprehensive taxonomy of barriers and facilitators organized according to the Theoretical Domains Framework, providing a structured resource for intervention developers and researchers. The review will generate a conceptual model illustrating pathways through which social isolation influences physical activity participation, identifying both direct effects and indirect influences mediated through psychological, social, and environmental factors. Evidence-based recommendations will be developed for programme design features that address isolation-specific barriers, implementation strategies for reaching and engaging socially isolated older adults, and evaluation approaches that capture both physical activity and social outcomes. The identification of evidence gaps will establish priorities for future research, including understudied populations, programme types, and contextual factors requiring further investigation.

The methodological strengths of this review include its comprehensive search strategy without language restrictions, systematic application of an established theoretical framework, rigorous quality assessment using validated tools, and transparent synthesis approach with clear audit trails. The use of Covidence software ensures systematic management of the review process and facilitates collaboration between reviewers while maintaining independence in screening and quality assessment decisions. The GRADE-CERQual assessment will provide transparent evaluation of confidence in review findings, enabling users to assess the strength of evidence supporting different recommendations. The involvement of multiple reviewers in all stages of the review process enhances reliability and reduces potential bias in study selection, quality assessment, and data synthesis.

The anticipated limitations of this review must be acknowledged to appropriately contextualize findings and recommendations. The heterogeneity of included studies in terms of populations, settings, programme types, and methodological approaches may complicate synthesis and limit the specificity of recommendations for particular contexts or subgroups. The broad definition of social isolation necessary to capture diverse conceptualizations may obscure important distinctions between different types and degrees of isolation that have differential impacts on participation. The focus on structured physical activity programmes may exclude important insights from studies examining informal or self-directed physical activity that could inform programme development. The synthesis of qualitative findings inherently involves interpretation, and despite efforts to maintain transparency and rigor, the review findings will inevitably be influenced by the reviewers’ perspectives and assumptions.

The potential challenges in conducting this review include the possibility of limited high-quality qualitative research specifically addressing social isolation and physical activity participation, requiring careful consideration of how to synthesize findings from studies where social isolation is a secondary focus. The complexity of the phenomenon under investigation, involving multiple interacting factors across different levels of influence, may make it difficult to develop clear, actionable recommendations that adequately account for contextual variation. The rapid evolution of physical activity programme delivery, particularly regarding technology-mediated approaches accelerated by the COVID-19 pandemic, means that review findings may need regular updating to maintain relevance. The translation of qualitative findings into practical recommendations requires careful attention to avoid oversimplification while maintaining utility for programme developers and policymakers.

## Data Availability

All data produced in the present study are available upon reasonable request to the authors.

